# The “Sensory Paradox”: Exploring the Positive Association Between Hyper- and Hypo-Responsivity to Sensory Stimuli in Autism and Beyond

**DOI:** 10.1101/2025.09.21.25336283

**Authors:** Kyle E. Takach, Kacie Dunham-Carr, Gerardo Parra, Linnea Joffe-Nelson, Lauren Jones, Reanna Mankaryous, Savannah Rogers, Catherine Serianni, Meiwen Shao, Bo Zhang, Ellen Hanson, Nicolaas A. Puts, Laura Cornelissen, April R. Levin

**Affiliations:** Department of Neurology, Boston Children’s Hospital, Harvard Medical School, Boston, MA, USA; Division of Developmental Medicine, Boston Children’s Hospital, Harvard Medical School, Boston, MA, USA; Biostatistics and Research Design Center, Institutional Centers for Clinical and Translational Research, Boston Children’s Hospital, Harvard Medical School, Boston, MA, USA; Department of Forensic and Neurodevelopmental Sciences, Institute of Psychiatry, Psychology, and Neuroscience, King’s College London, London, UK; MRC Centre for Neurodevelopmental Disorders, King’s College London, London, UK; Department of Anesthesiology, Critical Care and Pain Medicine, Boston Children’s Hospital, Harvard Medical School, Boston, MA, USA

**Keywords:** Autism, sensory processing, neurodevelopment

## Abstract

**Background:** Differences in sensory processing are a core feature of autism spectrum disorder. Hyper- and hyporesponsivity to sensory stimuli have historically been conceptualized as separate constructs but may co-occur within individuals. Sensory processing may impact both lower and higher-level cognitive processes; thus, it is crucial to understand the relationships between hyper- and hyporesponsivity within and across modalities, as well as the relationship between sensory processing and other aspects of development in both autistic and typically developing (TD) children.

**Methods:** In 3–4-year-old children (*n*=41 autism; *n*=37 TD), we assessed relationships between sensory hyper- and hyporesponsivity both within and across visual, auditory, touch, and oral sensory modalities as measured by caregiver report. Secondary analyses evaluated relationships between sensory responsivity, social communication, and cognitive abilities.

**Findings:** We found a positive correlation between sensory hyper- and hyporesponsivity (ρ = .788, *p* < .001). These associations persisted within groups and within and across modalities. There are positive associations between sensory responsivity and social interaction, communication, and nonverbal developmental quotient, with associations between sensory responsivity and social communication driven by associations within the autism group.

**Interpretation:** The positive correlations between hyper- and hyporesponsivity both within and across sensory modalities, which we term the “Sensory Paradox,” may provide key clues to understanding sensory processing in autism and other neurodevelopmental disorders by pointing towards neural circuit-level mechanisms that may underlie neurodevelopmental conditions.

**Funding:** This study was funded by NIH/NINDS 1R01NS134948-01 (ARL), NIMH T32MH112510 (KDC), the Simons Foundation Autism Research Initiative (Award number 648277, ARL), and the Eagles Autism Foundation (ARL).

**Research in context:** *Evidence before this study:* Up to 95% of autistic individuals are impacted by sensory processing differences. Across the full range of the autism spectrum, including individuals with profound ASD and self-advocates who speak publicly on issues of neurodiversity, improving sensory processing challenges is repeatedly noted as a common goal that would improve quality of life. Classical medical evaluation of sensory processing typically focuses on whether the structural pathways for transmission of sensory information are intact. The modulation of sensory information as it traverses these pathways, however, is a field ripe for further understanding. Initial reports have identified both hyper- and hyporesponsivity to sensory stimuli in autism, with some overlap between the two patterns of behavior.

*Added value of this study:* This study demonstrates the seemingly paradoxical finding that hyper- and hyporesponsivity are strongly positively correlated in both autistic and typically developing toddlers. This positive correlation persists within groups and within individual sensory modalities (sight, sound, touch, and oral), as well as across modalities.

*Implications of all of the available evidence:* The current findings, taken together with prior literature, support the Sensory Paradox - a framework for understanding sensory processing and the resulting sensory experience of autistic individuals which may also have key implications for a wider variety of neurological, psychiatric, and developmental conditions. Rather than considering hyper- and hyporesponsivity as static and opposing constructs, future work on the neurobiology, diagnosis, and management of sensory processing will benefit from considering the variable and context-dependent nature of sensory processing within individuals.

## Introduction

Differences in sensory processing are highly prevalent in autism spectrum disorder (henceforth referred to as “autism”, with those diagnosed referred to as “autistic”, per the preferences of the wider autism community^1^), impacting up to 95% of autistic individuals,^2–4^ a rate significantly higher than that found in typically developing (TD) individuals.^5, 6^

Sensory processing refers to how the nervous system registers, organizes, prioritizes, and interprets stimuli from the environment. Characteristics of the sensory input in a given modality (e.g., visual, auditory, tactile) are extensively processed through the hierarchal organization of the nervous system: for example, in response to a tap on the finger, a peripheral neuron sends the signal to neurons in the spinal cord, up through the thalamus, and into primary and integrative areas of cerebral cortex.^7^ At each level, from peripheral to central,^8^ opportunities for autoregulation, or self-regulation of neuronal firing,^9^ through bottom-up/top-down, feedforward/feedback/lateral, and excitation/inhibition/disinhibition interactions^10^ modulate the strength of this incoming signal.^11^ Therefore, the conscious perception of sensory stimuli reflects not only the characteristics of the sensory stimulus itself but also the nervous system’s modulation thereof. This modulation ultimately determines an individual’s perceptual sensitivity, physiological and affective reactivity, and resulting behavioral responsivity to sensory input.^12^

In much of prior research hyper- and hyporesponsivity are purportedly opposites – i.e., as if decreasing responsivity would be most helpful to those with hyperresponsivity and vice versa. These prior studies generally conceptualized hyper- and hypo-responsivity as separate, distinct constructs of sensory processing; this conceptualization is clinically useful and intuitive.

However, an increasing number of studies have noted both hyper- and hyporesponsivity to occur within and across different sensation modalities (e.g., visual, auditory, touch) in autism, even within the same individual.^2, 5, 13, 14^ This leads to an alternative conceptualization, setting the stage for considering how dynamic, moment-to-moment changes in neural circuit activity and environmental context could lead to variably hyper- or hypo-responsivity to sensory stimuli within a single individual, and possibly even within a single sensory modality.

Prior studies have demonstrated that sensory processing differences can be measured at multiple levels, from neural responses to sensation to behavior,^12^ and are associated with other core features of autism, including social communication and restricted, repetitive behaviors, and higher order cognitive processes.^15–17^ These associations are important because they suggest that sensory processing, in addition to directly impacting an individual’s experience of the world and quality of life, may have downstream effects on aspects of higher-level of cognition (and vice versa). Furthermore, sensory processing research broadly offers opportunities for translation across species, and thus a tractable window into the neurobiological and circuit-level mechanisms (and heterogeneity thereof) of higher-level social and cognitive function in ASD and related neurodevelopmental conditions.^11^

Sensory differences in autism arise early in life^18^ and persist across time.^3, 19^ However, a meta-analysis of sensory behaviors reveals non-linear developmental patterns, suggesting that age may contribute to variability of findings across studies,^13^ especially given that many prior studies of behavioral responses to sensory stimuli in autistic children focused on a fairly wide age range.^6^

In the current study we thus introduce, test, and consider the concept of the “Sensory Paradox;” i.e., the co-occurrence of hyper- and hyporesponsivity to sensory stimuli, both across and within modalities, within individuals. We focus on a small age range (3-4 years), near the time when children are old enough that behavioral manifestations of autism are generally recognizable and autism diagnosis tends to be relatively stable,^20^ but young enough that they are still early in their developmental trajectory and thus particularly responsive to therapeutic interventions. Through parent report questionnaires to assess sensory responsivity and core autism symptoms, alongside clinician observation to assess developmental level and early cognitive ability, we aim to answer the following questions in this age group:

1. What associations are present between levels of sensory responsivity (hyperresponsivity, hyporesponsivity) and various sensory modalities (vision, auditory, touch, oral)?
2. How is sensory processing associated with social communication, repetitive behavior, and cognition?

We hypothesized that sensory responsivity would be positively correlated across modalities (e.g., audition, vision) and domains (i.e., hyperresponsivity and hyporesponsivity), positively correlated with social communication, and negatively correlated with cognition.

## Methods

### Participants

78 participants (37 TD and 41 autism) between the ages of 36 and 59 months were recruited from local clinics, a Research Participant Registry (RPR), community flyers and tabling events. Participants with autism were also recruited using SPARK.^21^ The study was approved by the Institutional Review Board at Boston Children’s Hospital.

Participants included in the autism group had a community diagnosis of autism and met criteria for autism on the Autism Diagnostic Observation Schedule – 2^nd^ edition (ADOS-2^22^) administered by a research reliable staff member during the study visit. TD participants had no concerns for autism or other neurodevelopmental disorders, and no first degree relative with autism per parent report. For both groups, exclusion criteria also included known uncorrected vision or hearing impairments, motor disabilities that would preclude participation in testing, and use of medications known to affect sleep or cognition.

### Measures

#### Sensory Profile-2 (SP2)

The Sensory Profile-2 (SP2) is a caregiver report measure of behavioral reactions to sensory stimuli that are found throughout everyday life^23^ We analyzed total modality scores for auditory, visual, tactile, and oral sensory responsivity, along with the quadrant scores of Avoiding, Sensitivity, Registration, and Seeking. Following methodology from past studies,^5^ hyperresponsivity was measured by summing scores in the Sensitivity and Avoiding categories of the SP2 (e.g., “Is more bothered by bright lights than other same-aged children”; “Holds hands over ears to protect them from sound”), and hyporesponsivity was measured by summing scores in the Registration and Seeking categories (e.g., “Seems unaware of pain;” Displays need to touch toys, surfaces, or textures”).

#### Social Communication Questionnaire (SCQ)

The SCQ is a caregiver report asking about symptoms of autism including social communication and repetitive behavior.^24^ Higher scores indicate greater concern for autism, and as a screener, the “probable autism” cut-off for the SCQ is 12. Total scores, as well as raw totals from the Reciprocal Social Interaction, Restrictive, Repetitive, and Stereotyped Social Behaviors, and Communication Sum subscales, were derived for analyses.

#### Mullen Scales of Early Learning (MSEL)

Children in both autism and TD groups were administered the MSEL, a standardized developmental assessment for children from birth through 68 months of age.^25^ Four of the five MSEL subscales were administered: Visual Reception (VR), Fine Motor (FM), Expressive Language (EL), and Receptive Language (RL). Gross motor was not administered because all participants are above 36 months, which is more than the max age for the gross motor subscale. Standardized MSEL scores and age equivalents were derived for each subtest. The administered subscales allowed calculation of a verbal and non-verbal developmental quotient (DQ) for each participant.

### Statistical Analysis

Continuous variables were evaluated for normality, here defined as skewness <|1.0| and kurtosis <|3.0|. The visual and oral total domain scores from the SP2 and the Reciprocal Social Interaction and Restricted, Repetitive, and Stereotyped Patterns of Behavior subscales on the SCQ were transformed via square root transformation to correct for positive skew prior to analyses. The Sensitivity quadrant score and the hyperresponsivity aggregate score from the SP2 were unable to be transformed; thus, nonparametric analyses were used with these variables. Statistical analyses were performed in R.^26^ There was no missing data in the present sample.

TD and autism groups were compared on demographic variables and all dependent variables of interest using independent two-sample t-tests or Wilcoxon rank-sum tests for continuous variables, and chi-square tests for categorical variables. To answer our first research question, we conducted a series of correlation and regression analyses of hyper- and hyporesponsivivity aggregate scores, SP2 quadrant scores, and auditory, visual, tactile, and oral modalities. To answer our second research question, we conducted additional Pearson or Spearman correlation and regression analyses between hyper- and hyporesponsivity aggregate scores on SCQ and MSEL scores. A deliberate choice was made to not correct for multiple comparisons because this was an exploratory analysis.^27^

## Results

### Group Differences in Demographics, Clinical Characteristics, and SP2 Scores

Table 1 describes the demographic characteristics of the sample. Groups significantly differed in the proportion of males sampled (*p* = .014). Annual income also significantly differed between the TD and autism groups (*p* = .001).

**Table 1.** Demographic Characteristics of the TD and Autism cohorts.

| Demographic |  | TD | Autism | p-value |
| --- | --- | --- | --- | --- |
|  | Age in Years, *M* (SD) |  |  |  |
| Sex | Male | 48.6%<br>*n*=18 | 78.0%<br>*n*=32 | 0.014* |
|  | Female | 51.4%<br>*n*=19 | 21.95%<br>*n*=9 |  |
| Race | Asian | 5.41%<br>*n*=2 | 14.6%<br>*n*=6 | 0.198 |
|  | Black and/or African American | 8.11%<br>*n*=3 | 17.1%<br>*n*=7 |  |
|  | White | 78.4%<br>*n*=29 | 53.7%<br>*n*=22 |  |
|  | Multiracial | 8.11%<br>*n*=3 | 12.2%<br>*n*=5 |  |
|  | Other | 0%<br>*n*=0 | 2.44%<br>*n*=1 |  |
| Ethnicity | Hispanic | 13.5%<br>*n*=5 | 24.4%<br>*n*=10 | 0.353 |
|  | Not Hispanic | 86.5%<br>*n*=32 | 75.6%<br>*n*=39 |  |
| Annual Income | Less than \$80,000 | 5.41%<br>*n*=2 | 29.3%<br>*n*=12 | 0.001** |
| | \$80,000 to \$140,000 | 16.2%<br>*n*=6 | 39.0%<br>*n*=16 | |
| | \$140,000 or Greater | 73.0%<br>*n*=27 | 26.8%<br>*n*=11 | |
|  | No Response | 5.41%<br>*n*=2 | 4.88%<br>*n*=2 |  |

Except for age, all values are represented as counts and percentages.
\* $p < 0.05$ , \*\* $p < .001$

Table 2 details the clinical characteristics of the sample, to determine whether our sample replicates prior findings that autistic children differ in sensory responsivity from TD.^6^ Groups significantly differed on all Sensory Profile Quadrant scores, as well as auditory, visual, tactile and oral subscale total scores. Groups also significantly differed in aggregate hyperresponsivity (i.e., combined Sensitivity and Avoiding quadrant scores), *U* = 134, *p* <.001, as well as hyporesponsivity (i.e., combined Registration and Seeking quadrant scores), *t* = −7.15, *p* <.001 (Figure 1).

**Figure 1.**
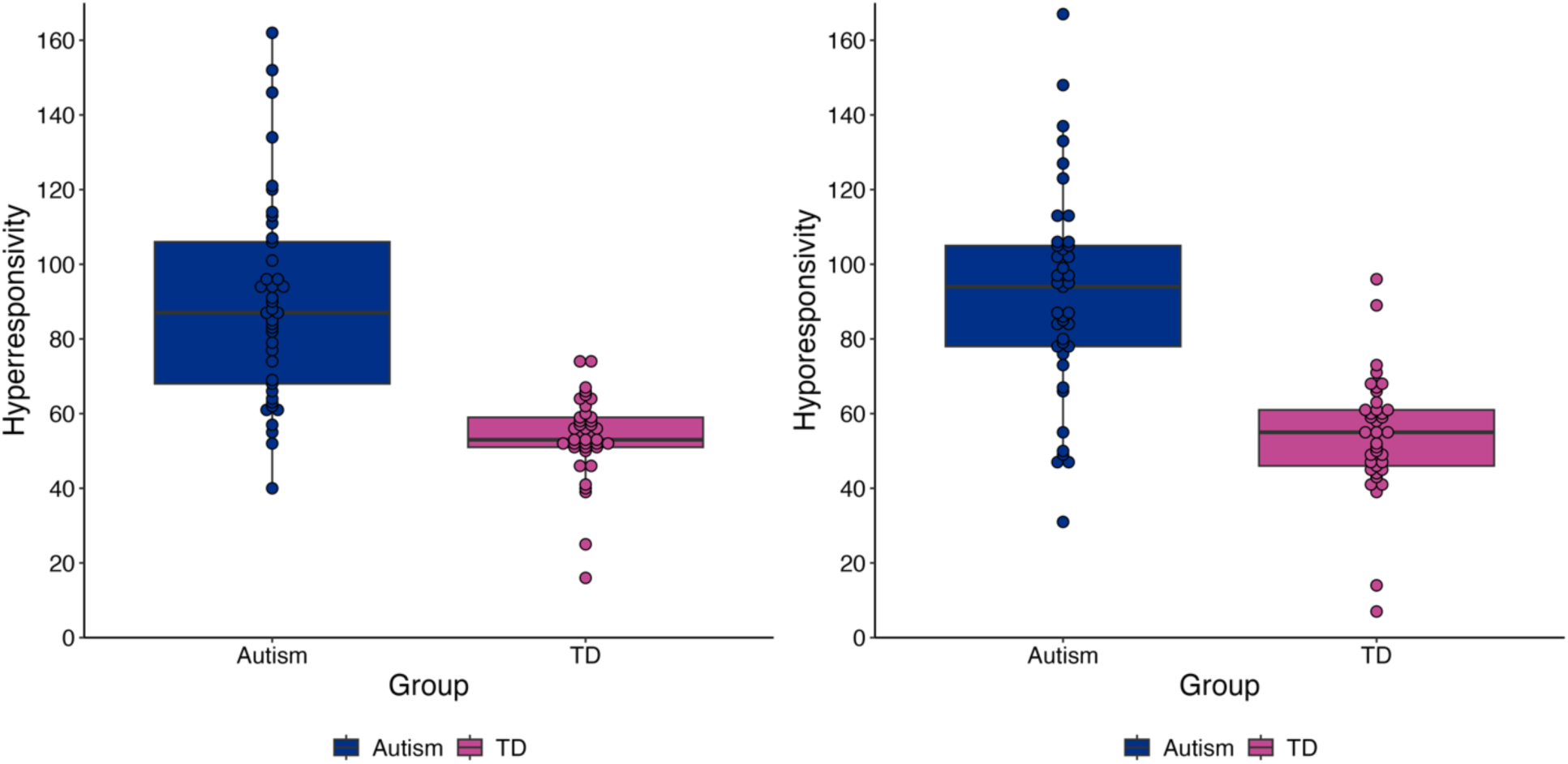
Total hyper- and hyporesponsivity scores across all modalities on the SP2 for each group. Blue = autism group, pink = typically developing group. TD = typically developing group.

**Table 2.** Sensory Responsivity, Autism Features and Developmental Quotients for TD and Autistic participants.

| Variable | TD<br>*M* (SD) | Autism<br>*M* (SD) | *p* value | Effect Size |
| --- | --- | --- | --- | --- |
| Hyperresponsivity | 53.7 (11.4) | 89.9 (27.7) | <.001*** | 0.708 |
| Hyporesponsivity | 54.5 (16.4) | 91.6 (28.4) | <.001*** | 1.58 |
| SP2 Seeking<br>Quadrant Score | 28.4 (8.57) | 47.1 (15.6) | <.001*** | 1.467 |
| SP2 Avoiding<br>Quadrant Score | 29.4 (7.17) | 42.9 (15.7) | <.001*** | 1.09 |
| SP2 Registration<br>Quadrant Score | 26.1 (9.13) | 44.6 (14.4) | <.001*** | 1.52 |
| SP2 Sensitivity<br>Quadrant Score | 24.3 (5.91) | 47.0 (13.8) | <.001*** | 0.811 |
| SP2 Auditory<br>Processing Total | 14.8 (5.28) | 22.1 (6.48) | <.001*** | 1.21 |
| SP2 Visual<br>Processing Total | 9.78 (2.99) | 11.9 (4.17) | .013* | 0.579 |
| SP2 Tactile<br>Processing Total | 12.4 (3.69) | 21.1 (7.72) | <.001*** | 1.42 |
| SP2 Oral Processing<br>Total | 13.7 (5.46) | 24.5 (11.5) | <.001*** | 1.14 |
| SCQ Total Score | 2.3 (1.81) | 17.9 (5.05) | <.001*** | 4.03 |
| SCQ Reciprocal<br>Social Interaction<br>Score | 0.76 (0.93) | 6.85 (3.40) | <.001*** | 2.62 |
| SCQ Restrictive,<br>Repetitive, and<br>Stereotyped<br>Behaviors Score | 0.84 (1.28) | 4.61 (1.73) | <.001*** | 2.60 |
| SCQ Communication<br>Sum | 1.68 (1.25) | 5.80 (1.83) | <.001*** | 2.61 |
| MSEL VQ | 121.5 (16.6) | 63.5 (39.0) | <.001*** | 1.90 |
| MSEL NVQ | 110.4 (16.8) | 66.3 (25.0) | <.001*** | 2.05 |

*Note.* Mean and standard deviation (SD) of scores are presented for each cohort. Effect sizes are Cohen's *d* for independent samples *t*-tests and the Wilcoxon *r* effect size for Wilcoxon rank-sum tests. SP2 = Sensory Profile 2, SCQ = Social Communication Questionnaire, MSEL = Mullen Scales of Early Learning.
\* $p < 0.05$ ; \*\* $p < 0.01$ , \*\*\* $p < .001$ .

Groups also significantly differed on SCQ total and Reciprocal Social Interaction, Restrictive, Repetitive, and Stereotyped Patterns of Behaviors, and Communication Sum subscales. The average SCQ total score for the autism group was 17.9, above the “probable autism” screening cutoff of 12, with scores ranging from 6 – 25. The average SCQ total score for the TD group was 2.3, well below the “probable autism” screening cutoff of 12, with scores ranging from 0 – 7. Finally, groups significantly differed on verbal and nonverbal developmental quotients derived from the MSEL.

### Relationships Between Hyper- and Hyporesponsivity Across and Within Domains

Across all modalities and groups, aggregate hyperresponsivity was significantly positively correlated with aggregate hyporesponsivity, ρ = .788, *p* < .001 (Figure 2). While group was a significant covariate in these associations, α = .564, *p* = .037, within-group correlations between aggregate hyper- and hyporesponsivity were significantly positive for both TD (ρ = .530, *p* <.001) and autism (ρ = .631, *p* < .001) groups. SP2 quadrant scores were all highly intercorrelated across groups (*p* values for all correlations <.001). Table 3 summarizes relations between quadrant scores across groups. Table 4 summarizes relations between hyper- and hyporesponsivity scores within modality. These scores were all significantly correlated to one another across groups and were not moderated by group.

**Figure 2.**
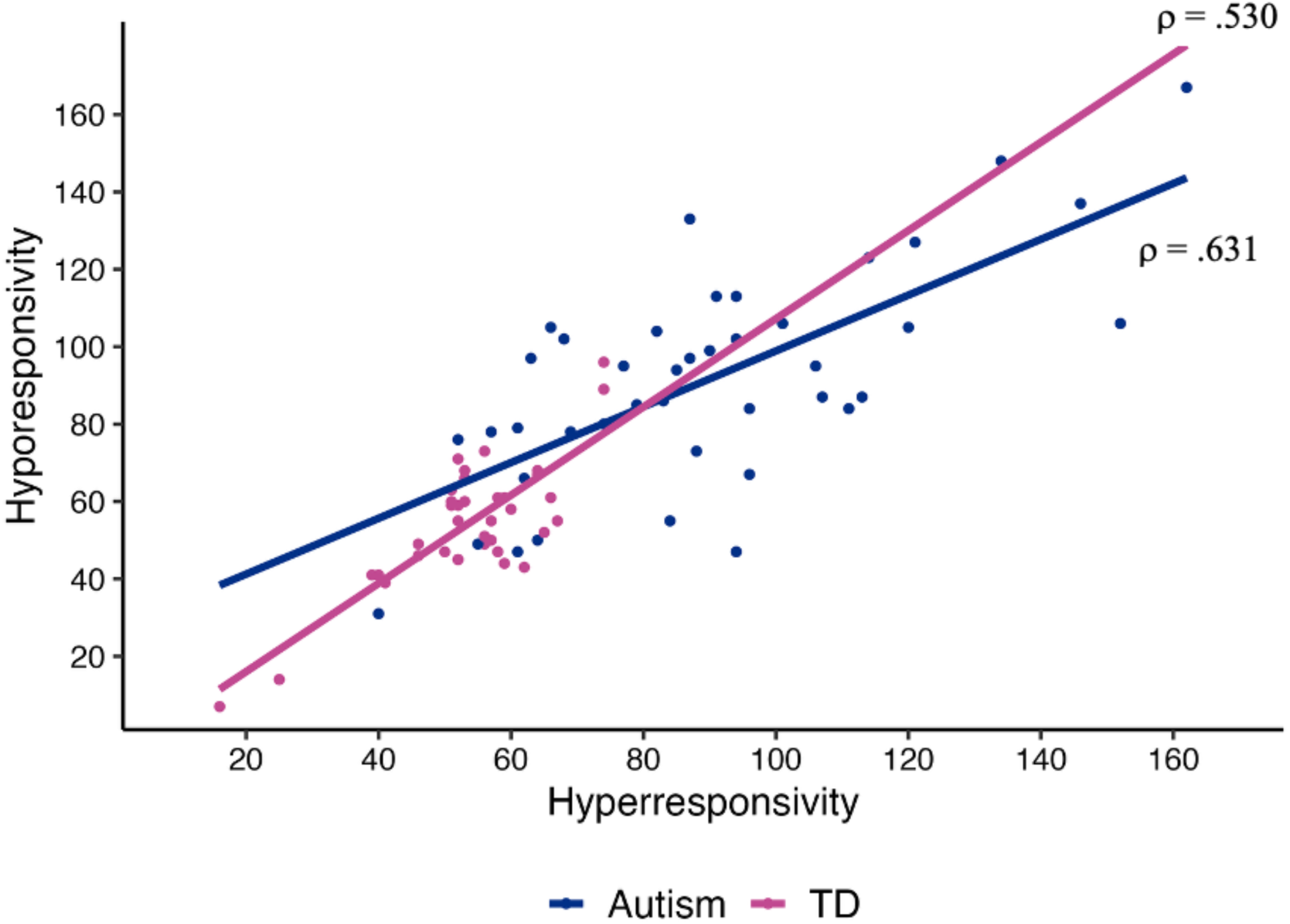
Correlation between total hyper- and hyporesponsivity scores across all modalities. Blue = autism group, Pink = typically developing group. TD = typically developing group.

**Table 3.** Correlations Between Sensory Domains.

| Domain | Registration | Seeking | Avoiding |
| --- | --- | --- | --- |
| Seeking | 0.861*** | - | - |
| Avoiding | 0.693*** | 0.786*** | - |
| Sensitivity | $\rho = 0.754^{***}$ | $\rho = 0.726^{***}$ | $\rho = 0.691^{***}$ |

*Note.* Correlation coefficients between each modality/domain are reported, unless otherwise specified as Spearman's rho.
\*\*\* $p < .001$ .

**Table 4.**
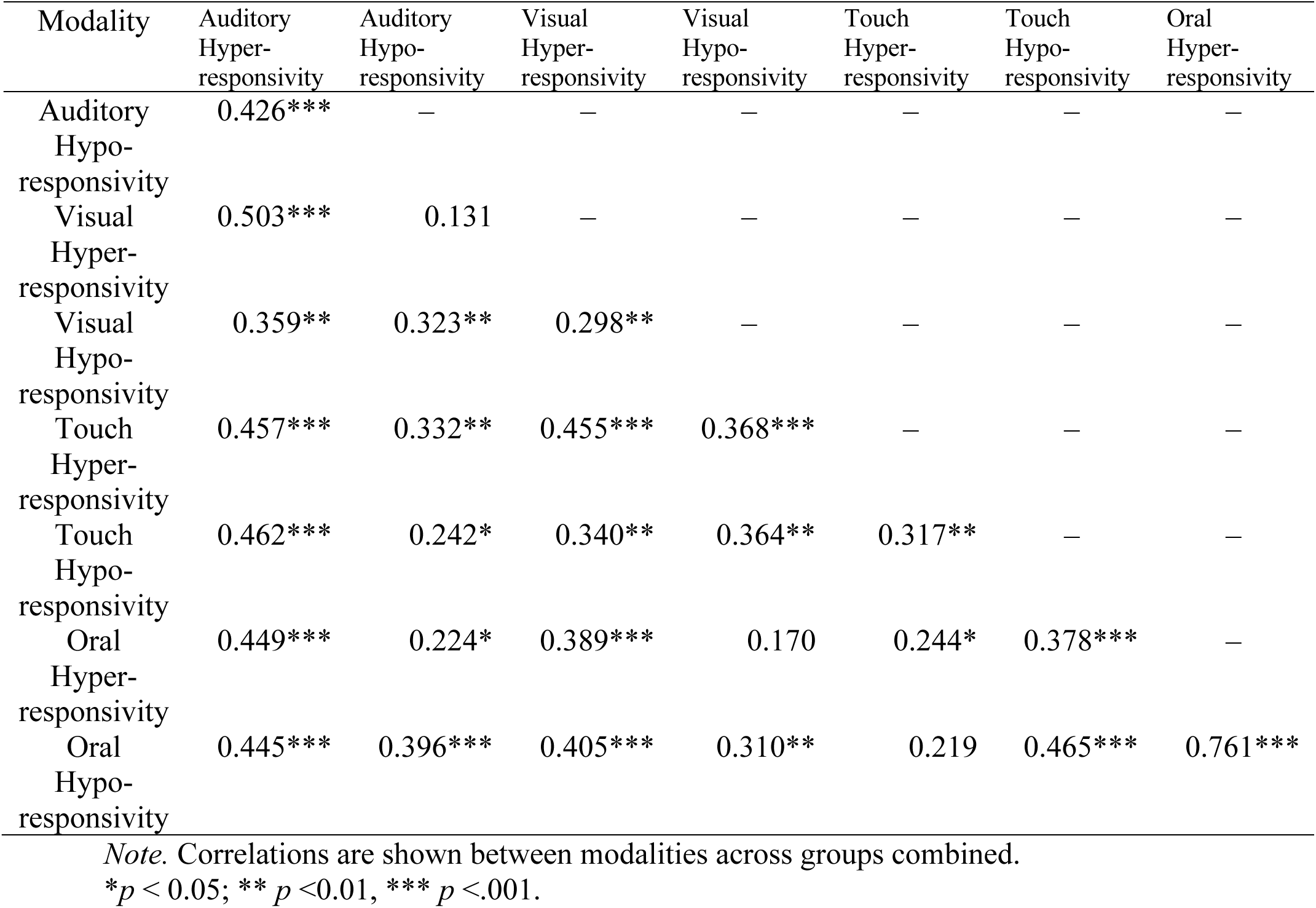
Correlations Within and Across Sensory Modalities.

### Relationships Between Sensory Responsivity, Autism Symptoms, and Developmental Quotients

Aggregate hyper- and hyporesponsivity scores were significantly correlated with SCQ Total scores, as well as with SCQ Communication Sum, SCQ Reciprocal Social Interaction, and SCQ Restrictive, Repetitive, and Stereotyped Behaviors (*p* values for all correlations <.001). Aggregate hyper- and hyporesponsivity scores were also significantly correlated with MSEL verbal developmental quotient and nonverbal developmental quotient (*p* values for all correlations <.001). Group was a significant covariate in associations between hyperresponsivity and SCQ Total scores and SCQ Communication Sum (*p* values for group term in regression models <.01), as well as in associations between hyporesponsivity and SCQ Total scores, SCQ Communication Sum, and SCQ Restrictive, Repetitive, and Stereotyped Behaviors (*p* values for group term in regression models <.05). Thus, within-group correlations between aggregate hyper- and hyporesponsivity and SCQ and MSEL metrics were run. From these within-group correlations, only associations between hyper- and hyporesponsivity and SCQ Total Scores and SCQ Restrictive, Repetitive, and Stereotyped Behaviors in the autism group and the association between hyperresponsivity and SCQ Communication Sum in the TD group remained significant. Table 5 summarizes associations between hyper- and hyporesponsivity and SCQ and MSEL metrics both within and across groups.

**Table 5.**
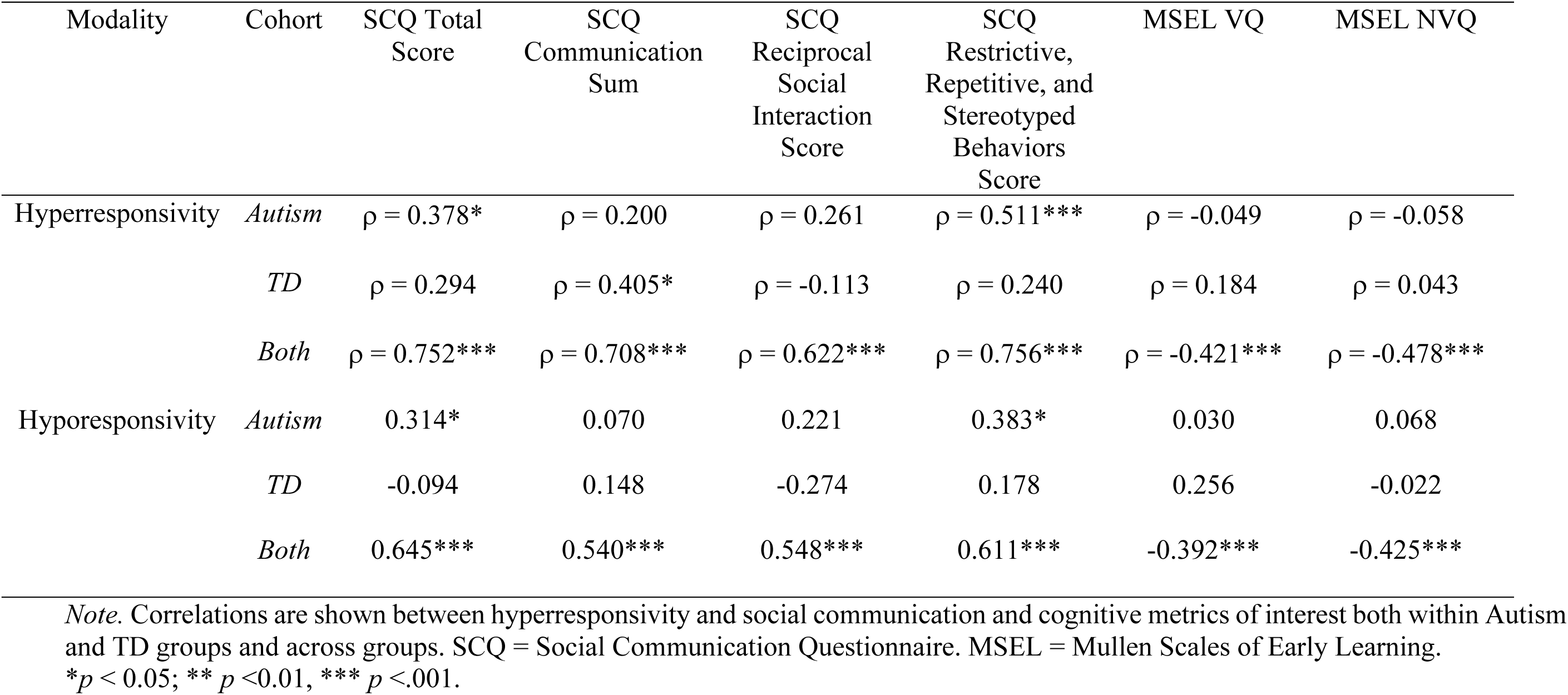
Correlations Between Hyper- and Hyporesponsivity and Social Communication and Cognitive Metrics.

| Modality | Cohort | SCQ Total<br>Score | SCQ<br>Communication<br>Sum | SCQ<br>Reciprocal<br>Social<br>Interaction<br>Score | SCQ<br>Restrictive,<br>Repetitive, and<br>Stereotyped<br>Behaviors<br>Score | MSEL VQ | MSEL NVQ |
| --- | --- | --- | --- | --- | --- | --- | --- |
| Hyperresponsivity | *Autism* | $\rho = 0.378^*$ | $\rho = 0.200$ | $\rho = 0.261$ | $\rho = 0.511^{***}$ | $\rho = -0.049$ | $\rho = -0.058$ |
| | *TD* | $\rho = 0.294$ | $\rho = 0.405^*$ | $\rho = -0.113$ | $\rho = 0.240$ | $\rho = 0.184$ | $\rho = 0.043$ |
| | *Both* | $\rho = 0.752^{***}$ | $\rho = 0.708^{***}$ | $\rho = 0.622^{***}$ | $\rho = 0.756^{***}$ | $\rho = -0.421^{***}$ | $\rho = -0.478^{***}$ |
| Hyporesponsivity | *Autism* | 0.314* | 0.070 | 0.221 | 0.383* | 0.030 | 0.068 |
|  | *TD* | -0.094 | 0.148 | -0.274 | 0.178 | 0.256 | -0.022 |
|  | *Both* | 0.645*** | 0.540*** | 0.548*** | 0.611*** | -0.392*** | -0.425*** |

*Note.* Correlations are shown between hyperresponsivity and social communication and cognitive metrics of interest both within Autism and TD groups and across groups. SCQ = Social Communication Questionnaire. MSEL = Mullen Scales of Early Learning.
\* $p < 0.05$ ; \*\* $p < 0.01$ , \*\*\* $p < .001$ .

## Discussion

This study demonstrates that hyper- and hyporesponsivity are positively correlated. Notably, hyper- and hyporesponsivity are not only positively correlated across all modalities, but also positively correlated within modalities, suggesting that these co-occurring patterns of responsivity are consistent through sensory modalities, rather than due to patterns of hyporesponsivity in some modalities and hyperresponsivity in others. There are also positive associations between sensory responsivity and social interaction, communication, and nonverbal DQ.

The “Sensory Paradox” (i.e., the finding that hyper- and hyporesponsivity are positively correlated) is interesting to consider. One metaphor to consider this concept is of the brain’s regulation of sensory reactivity as a thermostat. A tightly regulated thermostat does not allow temperatures to drop too low or too high beyond the ideal temperature, whereas a loosely regulated thermostat may have higher and lower extreme temperatures, acting as a “see-saw” mechanism. In autistic children, more loosely controlled autoregulation can occur at multiple biological levels;^9^ this may also explain the increased intraindividual (trial-by-trial) variability in response to sensory stimuli within and across modalities in these individuals.^28^

The methods and results presented herein do not allow us to test what causes hyper- and hyporesponsivity to be positively correlated; however, the Sensory Paradox may provide a conceptual framework for considering prior findings and suggesting future work. Autism has been alternately described as disorder of gain control,^29^ prediction,^30^ attenuated Bayesian priors,^31^ divisive normalization,^32, 33^ and autoregulation;^9^ indeed, many of these theories were established and tested in the sensory domain. Despite the varying nomenclature, each of these theories implicates an altered transformation of sensory signals by neural circuits, depending on the characteristics of an incoming stimulus and the context in which this stimulus occurs rather than a static up- or down-regulation of neural responses. Sensory sensitivity is highly context-dependent, with responses that are variable, nonlinear, and typically dependent on recent (and perhaps concurrent) stimuli, rather than a consistent or linear pattern of responsivity.^11^ The Sensory Paradox offers an opportunity to reflect upon the granular neurobiological principles of how neural circuits modulate and transform sensory inputs to make meaning from the environment.

Notably, the positive correlation between hyper- and hyposensitivity is present separately in both the autism and TD groups, with both groups representing relatively diverse racial, ethnic, and socioeconomic backgrounds. This suggests that the correlation is not driven solely by the presence of both hyper- and hypo-responsivity in the autism group and lack thereof in the TD group. Instead, while the autism group does indeed show elevated hyper- and hyporesponsivity overall, this correlation occurs across a wide range of sensory responsivity levels – even in typical development. This is consistent with prior findings suggesting that paradoxical sensory responses are also not specific to autism; for example, they have been noted in pain disorders^34^ and attention deficit hyperactivity disorder.^35^

On a related note, our finding that sensory responsivity tends to be highly correlated across modalities suggest opportunities for back-translation to test the extent to which cellular and synaptic mechanisms of sensory processing may also be recapitulated across sensory modalities. Clinical work on sensory processing may also consider the concept that the Sensory Paradox can reflect variable and context-dependent modulation of incoming sensory signals, as opposed to a static hyperresponsivity in one modality and hyporesponsivity in another.

There were several associations between sensory responsivity and other phenotypic presentations in our cohorts. Aggregate hypo- and hyperresponsivity scores were associated with SCQ subscores and total scores, as well as MSEL verbal and nonverbal developmental quotients. One could consider these findings from both causal and mechanistic perspectives. From a causal perspective, early differences in sensory processing and perception have been theorized to cascade onto the development of higher-order skills, potentially leading to differences in cognition and communication.^16^, see ^36^ for a comprehensive review

A key limitation in the current study is that sensory responsivity is measured here by parent report (SP2), which evaluates behaviorally observed phenotypes. While highly relevant to daily functioning in autistic individuals, behavior may reflect differences in modulation of incoming sensory stimuli at any step of the neural hierarchy (or combination thereof), from initial transduction of the stimulus into a neural signal up through conscious perception and ultimately the resulting motor (behavioral) response.^12, 36^ Notably, a complex mix of hyper- and hyposensitivity have been previously described across a variety of sensory studies using different methodologies,^13^ and self-reported by autistic adults.^37^ Hyper- and hypoexcitability of neural networks in response to sensory stimuli may vary within an individual depending on the brain region studied^38^ as well as on the spatial^32^ and temporal^39^ context of a sensory stimulus.

In clinical practice, the structural pathways for transmission of sensory information are reasonably well understood and considered in a variety of conditions, including deafness, blindness, and various focal lesions of the peripheral nerves, spinal cord, and brain. The modulation of sensory information as it traverses these pathways, however, is a field ripe for further understanding. Future studies may help to elucidate the Sensory Paradox by measuring dynamic, moment-to-moment responses to sensory inputs across a variety of preclinical and clinical models and conditions. Such studies should consider how factors such as internal state, specific characteristics of a sensory stimulus (e.g., intensity and modality), context (both spatial and temporal), and developmental trajectory may determine how the nervous system modulates responses to incoming sensory stimuli. The Sensory Paradox thus provides a framework that is crucial for understanding sensory processing and the resulting sensory experience of autistic individuals and may also have key implications for a wide variety of neurological, psychiatric, and developmental conditions.

## Data Availability

De-identified data produced in the present study are available upon reasonable request to the authors.

## Funding Sources

This work was funded by grants from NIH/NINDS 1R01NS134948-01 (ARL), NIMH T32MH112510 (KDC), the Simons Foundation Autism Research Initiative (Award number 648277, ARL), and the Eagles Autism Foundation (ARL).

## Statements and Declarations

ARL has consulted for Deerfield Management Company, L.P. (Lab 1636) and Jaguar Gene Therapy. The authors otherwise have no competing interests to declare that are relevant to the content of this article.

The data published here are part of a larger study in which data collection is ongoing. Thus, de-identified data for this publication may be made available upon reasonable request to the author.

## Author contributions

ARL, LC, and BZ contributed to the study conception and design. Data collection and analysis were performed by KT, KD-C, GP, LJ-N, LJ, RM, SR, CS, and MS. The manuscript was drafted by KT and KD-C and all authors contributed to data interpretation, as well as reviewing and editing the manuscript. All authors read and approved the final manuscript. ARL supervised all aspects of the study.

## Notes

### Author Declarations

The Institutional Review Board at Boston Childrens Hospital gave ethical approval for this work.

